# Diagnostic Utility of Antigen Detection Rapid Diagnostic Tests for Covid- 19: A Systematic Review and Meta-Analysis

**DOI:** 10.1101/2021.04.02.21254714

**Authors:** Mina Ebrahimi, Narges Nazari Harmooshi, Fakher Rahim

## Abstract

**Background:** Early detection of coronavirus disease (COVID-19) infection to improve disease management, becomes the greatest challenge. Despite high sensitivity of RT-PCR, not only it was reported that 20-67% of infected patients have false negative results. Rapid diagnostic tests (RDTs) are widely used as a point-of-care test for SARS-CoV-2 detection in both pharyngeal and blood specimens. To be less time*-*consuming, not seem so costly, and requiring no special training make it more favorable, but the low sensitivity is the main limitation. Several reports indicated rapid test of blood and pharyngeal samples has the same sensitivity as the RT-PCR, but some reports have lower sensitivity especial in asymptomatic patients.

**Methods:** In the present survey, we investigate the eligible studies for sensitivity and specificity of rapid tests and explore the factors that influence the result to help better diagnose COVID-19 infection. 20 studies met the inclusion criteria, which impose 33 different tests.

**Results:** Our findings showed, type of sample, type of assay, time of sampling, and load of virus influence on sensitivity of RDTs.

**Conclusion:** This research extends our knowledge of how to improve the sensitivity of RDTs to better diagnose of infected patients to address the controlling COVID-19 pandemic.

## Introduction

Early detection of coronavirus disease (COVID-19) infection to improve disease management, becomes the greatest challenge. Real-time PCR (RT-PCR) was introduced as a gold standard test in the laboratory. Despite high sensitivity of RT-PCR, not only it was reported that 20-67% of infected patients have false negative results, but also RT-PCR cannot differentiate between infectious and non-infectious SARS-CoV-2 particles (1,2). Rapid diagnostic tests (RDTs) are widely used as a point-of-care test for SARS-CoV-2 detection in both pharyngeal and blood specimens. To be less time-consuming, not seem so costly, and requiring no special training make it more favorable, but the low sensitivity is the main limitation. Additional to the rapid antigen (Ag) test, the rapid antibody (Ab) test was considered as a timely point-of-care test to detect IgG and IgM Abs in blood, plasma, and serum of patient with COVID-19 (3,4). In a recent study Ricks *et al*. analyzed the health system cost and health impact of using RDTs among hospitalization and mildly symptomatic patient with COVID-19, and report that despite the low sensitivity of RDTs compare with RT-PCR, is accompanied with “Ag-RDTs have the potential to be simultaneously more impactful with a lower cost per death and infectious person-days averted” (5).

Several reports indicated rapid test of blood and pharyngeal samples has the same sensitivity as the RT-PCR, but some reports have lower sensitivity especial in asymptomatic patients (6–10). These controversial results may be due to the different sample types or examined in different infection stages. In the present systematic review, we attempt to assess the diagnostic utility of antigen detection rapid diagnostic tests for covid-19 versus RT-PCR in a different type of samples and different stages of infection determine the usability of rapid tests in best time and sample.

## Materials and Methods

This review was performed following the PRISMA (Preferred Reporting Items for Systematic reviews and Meta-Analyses) and MOOSE (Meta-analyses Of Observational Studies in Epidemiology) guidelines (11,12).

### Search Strategy

To evaluate the usability of rapid tests compare with RT-PCR, we systematically searched the electronic database, including Scopus, Medline/PubMed, EMBASE, Web of sciences (WOS), and Cochrane library using Mesh-standardized keywords: (((Rapid antigen detection test * OR RDT*) OR “Rapid Antigen Test ”[Mesh] OR “point of care testing”) AND (“Real-time PCR”[Mesh] OR “RT-PCR” OR “Molecular diagnostic test” OR “RNA virus”) AND (“2019 nCoV”[tiab] OR 2019nCoV[tiab] OR “2019 novel coronavirus”[tiab] OR “COVID 19”[tiab] OR COVID19[tiab] OR “new coronavirus”[tiab] OR “novel coronavirus”[tiab] OR “novel coronavirus”[tiab] OR “SARS CoV-2”[tiab] OR (Wuhan[tiab] AND (coronavirus[tiab] OR “corona virus”[tiab]) OR “COVID-19”[Supplementary Concept] OR “severe acute respiratory syndrome coronavirus 2”[Supplementary Concept])) until Jan 2021. There is no restriction for time and language, and the citation lists of selected articles were hand-searched for additional papers.

### Data extraction

Two reviewers (ME and FR) independently screened titles and abstracts of all initially found articles. Information was extracted from selected studies, including the name of the author, country, sample size, mean age, rapid diagnostic kits, true positive, false positive, false negative, true negative results, sensitivity, and specificity. A third reviewer was consulted to resolve any disagreements between reviewers by discussion until consensus was reached.

### Eligible Criteria

In order to understand the sensitivity and specificity of RDTs, studies that evaluated these parameters were selected. Inclusion criteria were considered as following: evaluation of the sensitivity and specificity of RDTs compare to the RT-PCR. All types of studies, including case/control, cohort, cross-sectional, and clinical trial studies, were included. Additionally, letter to editors which reported comprehensive data was included. Studies that evaluated seroprevalence, studies that investigated just cell culture assay, case reports, reviews, and studies reporting cases with incomplete information were excluded.

### Statistical Analysis

Cochran Chi-square test and *I*^*2*^ were used to assessing heterogeneity among studies. A fixed-effects model was used when *I*^*2*^ < 50%, while in the case of *I*^*2*^> 50%, a random-effects model was selected. Fixed-model assumes that the population effect sizes are the same for all studies (13). In contrast, the random-effects model attempted to generalize findings beyond the included studies by assuming that the selected studies are random samples from a larger population (14). To compare the sensitivity and specificity in RDTs compared with the RT-PCR, 95% confidence intervals (CI) were used. According to the heterogeneity test results, either Der Simonian’s and Laird’s random-effects method or Mantel-Haenszel’s fixed-effects method were used to estimate the overall sensitivity and specificity and 95% confidence intervals (15). Moreover, subgroup analysis was implemented based on the type of specimen (nasopharyngeal swab, throat washing and bronchoalveolar fluids, and Nasal sample), and symptomatic or asymptomatic patinates as an important variable which may cause heterogeneity between different samples or influence of onset of symptoms. The Egger’s test was used to investigate small study effects due to potential publication bias (16,17). If there was statistical heterogeneity among the results, a further sensitivity analysis was conducted to determine the source of heterogeneity. After the significant clinical heterogeneity was excluded, the randomized effects model was used for meta-analysis. P < 0.05 was considered as statistical significance (2-sided). All data were analyzed using STAT 16 (STATA Corporation, College Station, Texas).

## Result

Overall, 783 studies were initially collected. After removing duplicated studies, 580 studies have remained. During screening titles and abstracts, 165 studies were considered potentially eligible. Subsequently, in full-text screening, 295 studies were excluded (**Figure 1**). Finally, 20 studies met our inclusion criteria, which impose 33 different tests (including 26,056 patients, mea age range from 20.5 to 53.14 years). Eleven studies (55%) evaluated nasopharyngeal swabs (5,6,8,12–20). Three of included studies investigated various types of samples with just one assay (18,27,28), whereas other two articles examined one type of sample with different assays to comparing sensitivity and specificity (7,9). A single study performed a similar series of experiments but for rapid Abs tests with finger-stick whole-blood (10), whereas other two studies were used the same rapid Abs tests from patient’s serum (29,30). Characteristics of diagnostic values of included studies are shown in **Table 1**. Forest plots illustrating sensitivity and specificity for each analysis are seen in **Figure 2**. The results of Cochrane Q and *I*^*2*^ statistics showed significant heterogeneity in sensitivity and specificity, so estimations of sensitivity and specificity were obtained using a random effect model. In further analysis, data were analyzed based on the type of sample and symptomatic or asymptomatic patients **Table S1-S5** and **Figure S1-S5**. Summary ROC curves constructed for all assays based on Monte Carlo simulations are shown in **Figure 3**. It did not apply to assessing the false results, because 7 studies (35%) did not focus on false results (7,9,18,22,28,30–32). Some of the current kinds of literature which focused on Ab rapid tests were excluded due to reporting the results as the separate sensitivity for IgG and IgM or analyzed data based on onset of symptoms (3,33).

**Table 1.** Pooled analysis of sensitivity and specificity of included studies with 95% confidence interval.

| Study ID | Sensitivity | Specificity | Positive LR | Negative LR | DOR |
| --- | --- | --- | --- | --- | --- |
| Agullo et al.(27) | 0.576 (0.487- 0.661) | 0.998 (0.989-1.000) | 299.39 (42.023-2133.1) | 0.425 (0.348-0.519) | 704.36 (96.091-5163) |
| Agullo et al.(27) | 0.447 (0.360-0.536) | 1.000 (0.993-1.000) | 472.42 (29.398-7591.7) | 0.553 (0.475-0.645) | 854.05 (52.241-13962.1) |
| Agullo et al.(27) | 0.231 (0.160-0.317) | 1.000 (0.992-1.000) | 228.93 (14.076-3723.5) | 0.767 (0.696-0.846) | 298.41 (18.060-4930.7) |
| Agullo et al.(27) | 0.496 (0.404-0.588) | 1.000 (0.992-1.000) | 485.98 (30.265-7803.7) | 0.505 (0.423-0.602) | 963.08 (58.809-15771.9) |
| Abdelrazik et al. | 0.431 (0.359-0.505) | 1.000 (0.989-1.000) | 286.33 (17.860-4590.3) | 0.570 (0.503-0.645) | 502.65 (30.910-8173.8) |
| Albert et al. (21) | 0.796 (0.665-0.894) | 1.000 (0.990-1.000) | 567.87 (35.470-9091.7) | 0.209 (0.125-0.350) | 2712.1 (157.06-46833.5) |
| Ciotti et al.(6) | 0.308 (0.170-0.476) | 1.000 (0.715-1.000) | 7.500 (0.478-117.57) | 0.717 (0.564-0.912) | 10.455 (0.570-191.78) |
| Kohmer et al.(7) | 0.290 (0.204-0.389) | 0.250 (0.169-0.347) | 0.387 (0.279-0.536) | 2.840 (1.978-4.078) | 0.136 (0.073-0.255) |
| Kohmer et al.(7) | 0.320 (0.230-0.421) | 0.260 (0.177-0.357) | 0.432(0.318-0.589) | 2.615 (1.830-3.737) | 0.165 (0.090-0.305) |
| Kohmer et al.(7) | 0.180 (0.110-0.269) | 0.260 (0.177-0.357) | 0.243 (0.158-0.375) | 3.154 (2.238-4.445) | 0.077 (0.039-0.152) |
| Kohmer et al.(7) | 0.370 (0.276-0.472) | 0.260 (0.177-0.357) | 0.500 (0.378-0.662) | 2.423 (1.685-3.484) | 0.206 (0.113-0.377) |
| Linares et al. (22) | 0.157 (0.114-0.207) | 0.922 (0.881-0.951) | 2.000 (1.203-3.324) | 0.915 (0.858-0.975) | 2.186 (1.239-3.857) |
| Nalumansia et al.(23) | 0.700 (0.594 -0.792) | 0.924 (0.874-0.959) | 9.262 (5.398-15.891) | 0.325 (0.236-0.446) | 28.538 (13.848-58.814) |
| Pilarowski et al.(34) | 0.023 (0.008-0.053) | 0.960 (0.925-0.982) | 0.576 (0.196-1.691) | 1.018 (0.984-1.052) | 0.566 (0.187-1.717) |
| Pilarowski et al.(34) | 0.556 (0.212-0.863) | 0.503 (0.462-0.545) | 1.119 (0.620-2.018) | 0.883 (0.423-1.841) | 1.267 (0.337-4.767) |
| Salvagno et al.(31) | 0.340 (0.288-0.394) | 0.994 (0.978-0.999) | 54.500 (13.575-218.81) | 0.665 (0.614-0.719) | 82.007 (20.035-335.66) |
| Schildgen et al.(32) | 0.329 (0.223-0.449) | 0.877 (0.779-0.942) | 2.667 (1.332-5.338) | 0.766 (0.638-0.919) | 3.483 (1.486-8.162) |
| Schildgen et al.(32) | 0.500 (0.381-0.619) | 0.781 (0.669-0.869) | 2.281 (1.399-3.721) | 0.640 (0.495-0.829) | 3.563 (1.738-7.302) |
| Schildgen et al.(32) | 0.877 (0.779-0.942) | 0.795 (0.684-0.880) | 4.267 (2.696-6.753) | 0.155 (0.083-0.289) | 27.496 (11.184-67.599) |
| Scohy et al.(24) | 0.387 (0.291-0.472) | 1.000 (0.916-1.000) | 32.608 (2.053-517.87) | 0.628 (0.544-0.725) | 51.913 (3.118-864.30) |
| Toptan et al. (25) | 0.500 (0.319-0.681) | 1.000 (0.907-1.000) | 39.000 (2.432-625.53) | 0.506 (0.359-0.714) | 77.000 (4.357-1360.8) |
| Torres et al.(26) | 0.060 (0.043-0.081) | 1.000 (0.994-1.000) | 77.000 (4.741-1250.6) | 0.940 (0.922-0.959) | 81.905 (5.021-1336.2) |
| CK Mak et al.(18) | 0.343 (0.191-0.522) | 1.000 (0.900-1.000) | 25.000 (1.538-406.50) | 0.662 (0.520-0.843) | 37.766 (2.132-668.99) |
| CK Mak et al. (18) | 0.457 (0.288-0.634) | 1.000 (0.900-1.000) | 33.000 (2.057-529.44) | 0.549 (0.406-0.744) | 60.077 (3.416-1056.6) |
| CK Mak et al. (18) | 0.111 (0.037-0.241) | 1.000 (0.921-1.000) | 11.000 (0.626-193.25) | 0.890 (0.797-0.994) | 12.358 (0.663-230.48) |
| CK Mak et al. (18) | 0.400 (0.257-0.557) | 1.000 (0.921-1.000) | 37.000 (2.297-595.89) | 0.604 (0.476-0.768) | 61.218 (3.546-1056.9) |
| Prince-Guerra et al. (35) | 0.525 (0.467-0.583) | 0.999 (0.997-1.000) | 409.57 (152.91-1097.0) | 0.476 (0.422-0.536) | 861.29 (314.78-2356.6) |
| Courtellemont et al.(9) | 0.967 (0.918-0.991) | 1.000 (0.971-1.000) | 246.56 (15.502-3921.4) | 0.037 (0.015-0.092) | 6658.3 (354.65-125004.7) |
| Courtellemont et al.(9) | 0.706 (0.525-0.849) | 1.000 (0.897-1.000) | 49.000 (3.100-774.56) | 0.304 (0.183-0.506) | 161.0 (9.002-2879.3) |
| Pere et al. (10) | 0.958 (0.857-0.995) | 0.981 (0.897-1.000) | 49.833 (7.147-347.45) | 0.042 (0.011-0.165) | 1173.0 (102.92-13368.3) |
| Pere et al. (10) | 0.917 (0.800-0.977) | 0.865 (0.742-0.944) | 6.810 (3.401-13.636) | 0.096 (0.037-0.248) | 70.714 (19.333-258.66) |
| Pere et al. (10) | 0.923 (0.749-0.991) | 1.000 (0.858-1.000) | 45.37 (2.910-707.30) | 0.094 (0.029-0.308) | 480.20 (21.093-10527.9) |
| Pere et al.(10) | 0.979 (0.889-0.999) | 0.981 (0.897-1.000) | 50.917 (7.306-354.84) | 0.021 (0.003-0.148) | 2397.0 (145.76-39418) |
| Pere et al. (10) | 0.915 (0.796-0.976) | 0.846 (0.719-0.931) | 5.947 (3.125-11.316) | 0.101 (0.039-0.259) | 59.125 (16.576-210.89) |
| Fabre et al. (29) | 0.041 (0.017-0.083) | 0.959 (0.917-0.981) | 1.000 (0.359-2.789) | 1.000 (0.957-1.045) | 1.000 (0.343-2.916) |
| Cerutti et al.(36) | 0.706 (0.612-0.790) | 1.000 (0.983-1.000) | 312.82 (19.576-4998.8) | 0.296 (0.222-0.395) | 1056.4 (63.918-17459.1) |
| Montesinosa et al.(30) | 0.719 (0.632-0.795) | 1.000 (0.950-1.000) | 104.69 (6.597-1661.4) | 0.285 (0.216-0.375) | 367.47 (22.176-6089.1) |
| Montesinosa et al. (30) | 0.688 (0.600-0.766) | 0.958 (0.883-0.991) | 16.500 (5.417-50.263) | 0.326 (0.251-0.424) | 50.600 (15.016-170.51) |
| Montesinosa et al. (30) | 0.711 (0.624-0.788) | 1.000 (0.950-1.000) | 103.56 (6.525-1643.6) | 0.293 (0.223-0.384) | 353.80 (21.360-5860.2) |

**Figure 1.**
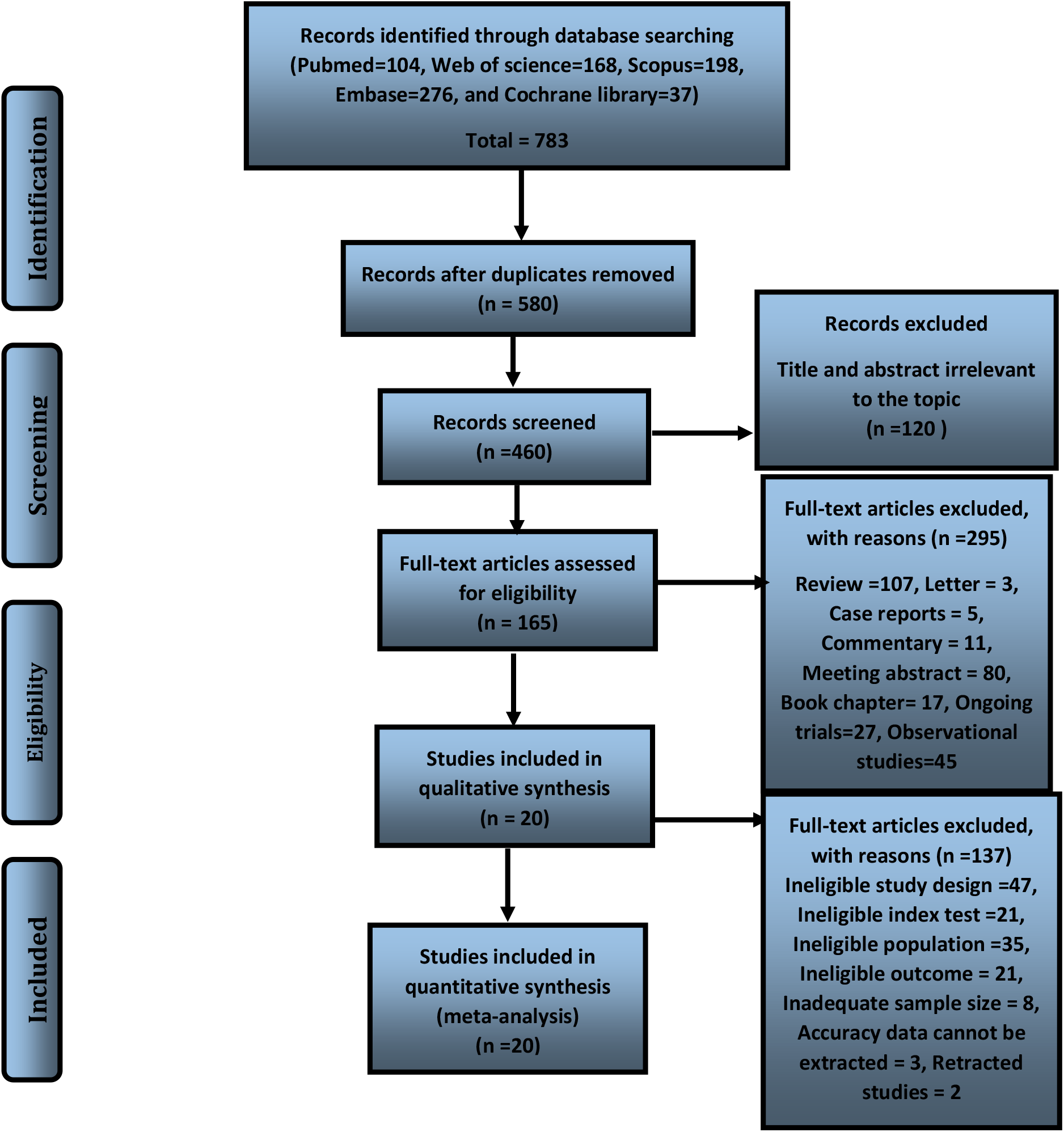
Flow diagram of the study selection process.

**Figure 2.**
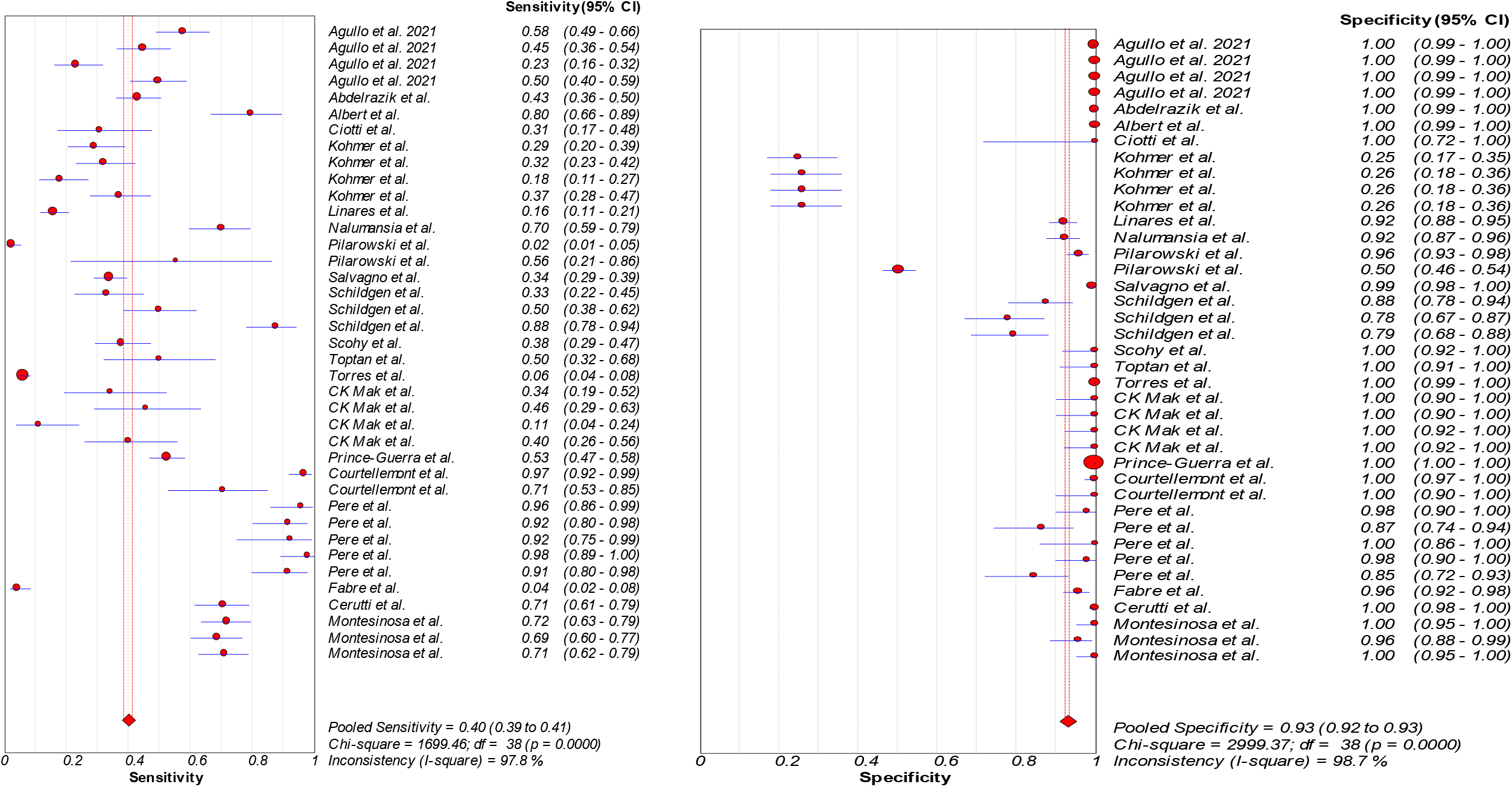
A forest plot showing the estimates for sensitivity (a) and specificity (b) as overall.

**Figure 3.**
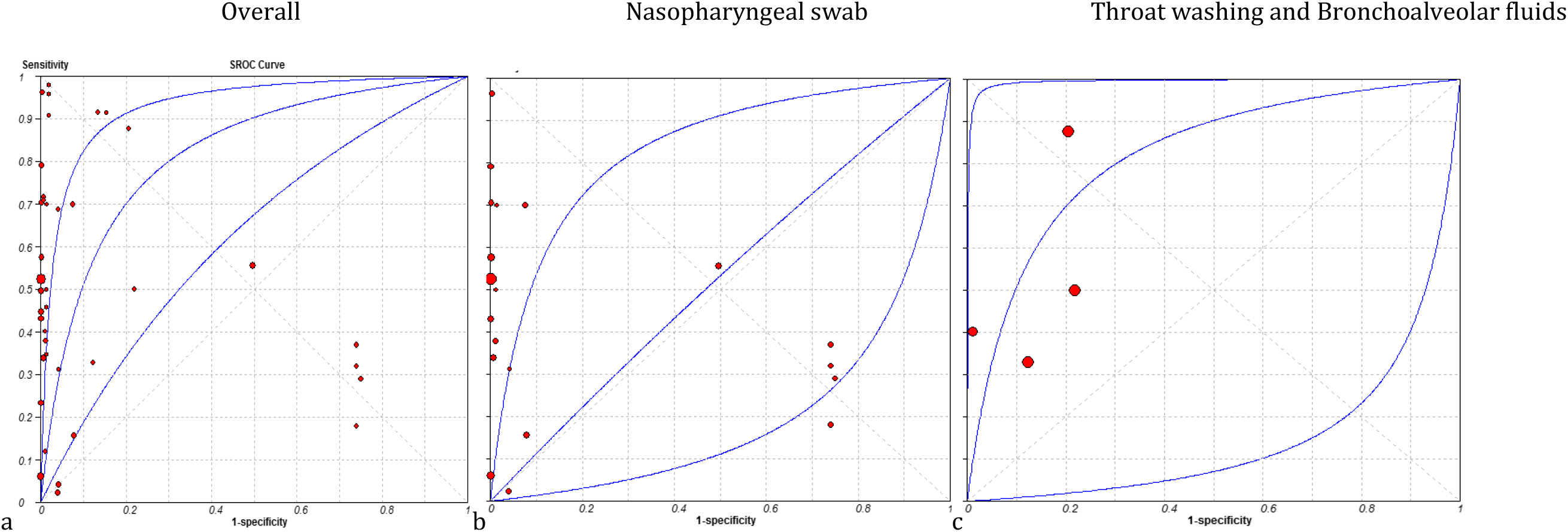
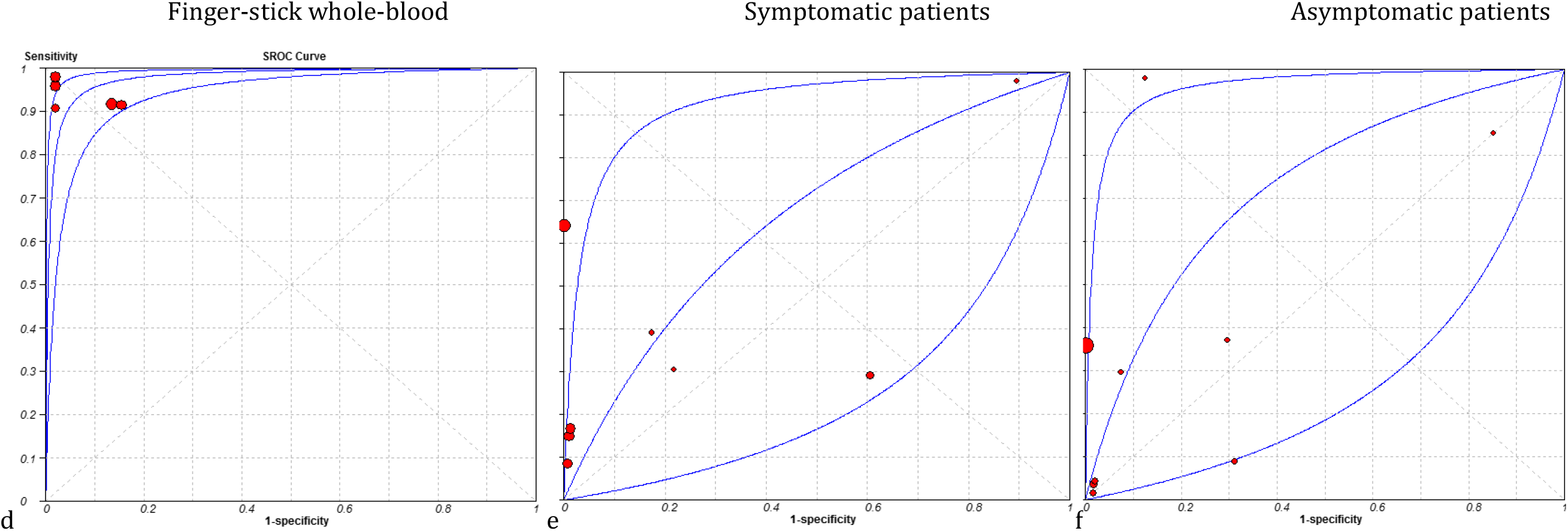
Summary receiver operating characteristic curve (ROC curve). Estimates of sensitivity and specificity for each study are

## Discussion

Following broad speared spectrum of COVID-19 infection, identified infected patients and carriers as soon as possible help to better control pandemic. In this line, the RDTs and rapid Abs test for detecting patients were introduced by WHO in a short time. A large and growing body of literature has investigated these tests’ usability based on their sensitivity and specificity compared to the RT-PCR. Inconsistency in reported results has heightened the need for a comprehensive search to achieving a reliable finding. In this regard, in the present survey, we investigate the eligible studies for sensitivity and specificity of rapid tests and explore the factors that influence the result to help better diagnose COVID-19 infection.

One important factor that influences the sensitivity of RDTs is a type of sample. Studies that evaluated different types of samples revealed that nasal examination swabs and saliva specimens for RDTs are associated with lower sensitivity (27). Our results showed the most collected samples were nasopharyngeal swabs; in this line, it was shown examine nasopharyngeal swab, the sensitivity of RDTs results increases (9,38). After evaluating various specimens, Yamayoshi *et al*. found that soaking specimen directly to the lysing buffer can improve RDTs sensitivity (38).

Our analysis has shown the higher sensitivity observes in infected patients with lower Ct value by RT-PCR. In fact, in asymptomatic patients or patients with lower viral load, the sensitivity of RDTs decreases (6,8,39). In the subsequent analysis, Cerutti et al. examine the STANDARD Q COVID-19 Ags (R-Ag) among infected patients; they demonstrated that patients who were positive for R-Ag have higher viral load and lower Ct value are symptomatic, and this can be an explanation higher RDTs sensitivity in symptomatic subjects (19). In contrast, Pilarowski *et al*. reported asymptomatic subjects should not rule out because regardless of symptoms, it can be possible asymptomatic patients have a viral load (40). In this regard, to the assessment of RDTs sensitivity, Eshghifar *et al*. used various concentration of heat-inactivated COVID-19 virus to assess a cut-off detection for RDTs; their findings showed, they could not determine a cut-off and reported RDTs to be positive just in patients with high viral load (28); whereas CK Mak *et al*. by using cell culture, the limiting of detection for RDTs reported 18.57 (18).

Additional to the RDTs, some writers compared the sensitivity of cell culture with RT-PCR for detecting COVID-19. Like the RDTs, the result showed that in cell culture with lowing viral load, the sensitivity reduces (8). This can be explained by the fact that specimens with low viral load or Ct value >30 do not cultured, suggesting low infectivity of virus (41).

Conducting RDTs with various assays is a serious discussion that can cause emerging different sensitivity. A study that set out to determine sensitivity of various assay for RDTs found different results for positive controls in different assays; which could offer the current methods detect different Ags and subsequently influence test accuracy (32).

The possible interference of time of testing cannot be ruled out. Some researchers revealed that testing less than 5 days from the onset of symptoms increases sensitivity (8,9,22,28). It is inconsistent with previous reports that a load of infectious virus decreases after 7-10 days (9,42). In one recent well-known recent experiment, Albert et al. with considering the age of participants found the sensitivity of RDTs is lower in pediatrics than adults (8).

Some recent attention has focused on the benefits of rapid Abs COVID-19 detection as one of the point-of-care tests. Our findings demonstrated that the rapid Ab detection has lower sensitivity than RDTs; it is accompanied by high false positives. Pere et al., in their investigation, showed a history of recent infection with cold Abs increase the false positive Ab rapid test, especially for IgM, which reduce the validation of the test (10). Fabre et al. performed a similar survey, but in pregnancy, surprisingly, their findings showed, the false positive is higher in pregnant women (29). One reason for the low accuracy of the rapid Ab test can be pointed out to the short-term immunity in such patients (43).

Much of the current systematic and meta-analysis literature focused on the accuracy of serological tests, and as the expected, their findings have shown lower accuracy of serological tests (44–46).

## Conclusion

The main goal of the current study was to determine the accuracy of RDTs. This study has found that generally, as the expected, RDTs have lower sensitivity than RT-PCR; this issue is affected by several factors such as type of specimen, the timing of sampling, type of assay, and viral load. This research extends our knowledge of how to improve the sensitivity of RDTs to better diagnose of infected patients to address the controlling COVID-19 pandemic.

## Data Availability

All data presented in the manuscript.

## Acknowledgment

No funding was received for the present study.

## Authors’ Contributions

F.R. conceived the manuscript and revised it. M.E and N.N done the statistical analysis and wrote the manuscript, and prepared tables and figures.

## Conflict of interest

The authors declare no conflict of interest. All procedure performs in studies involving human participants were in accordance with the ethical standards of the institutional and/or national research committee and with the 1964 Helsinki declaration and its later amendments or compare ethical strand.

## Notes

### Competing Interest Statement

The authors have declared no competing interest.

