## supplementary for "Diagnostic Utility of Antigen Detection Rapid Diagnostic Tests for Covid- 19: A Systematic Review and Meta-Analysis"

**Abstract**

**Background:** Early detection of coronavirus disease (COVID-19) infection to improve disease management, becomes the greatest challenge. Despite high sensitivity of RT-PCR, not only it was reported that 20-67% of infected patients have false negative results. Rapid diagnostic tests (RDTs) are widely used as a point-of-care test for SARS-CoV-2 detection in both pharyngeal and blood specimens. To be less time*-*consuming**,** not seem so costly, and requiring no special training make it more favorable, but the low sensitivity is the main limitation. Several reports indicated rapid test of blood and pharyngeal samples has the same sensitivity as the RT-PCR, but some reports have lower sensitivity especial in asymptomatic patients. **Methods:** In the present survey, we investigate the eligible studies for sensitivity and specificity of rapid tests and explore the factors that influence the result to help better diagnose COVID-19 infection. 20 studies met the inclusion criteria, which impose 33 different tests. **Results:** Our findings showed, type of sample, type of assay, time of sampling, and load of virus influence on sensitivity of RDTs. **Conclusion:** This research extends our knowledge of how to improve the sensitivity of RDTs to better diagnose of infected patients to address the controlling COVID-19 pandemic.

**Supplementary**

| Study | Sensitivity | Specificity | Positive LR | Negative LR | DOR |
| --- | --- | --- | --- | --- | --- |
| Agullo et al. (27) | 0.576 (0.487- 0.661) | 0.998 (0.989 - 1.000) | 299.39 (42.023 -2133.1) | 0.425 (0.348 - 0.519) | 704.36 (96.091 - 5163.0) |
| Abdelrazik et al. (37) | 0.431 (0.359- 0.505) | 1.000 (0.989 - 1.000) | 286.33 (17.860 -4590.3) | 0.570 (0.503 - 0.645) | 502.65(30.910 - 8173.8) |
| Albert et al. (21) | 0.796 (0.665- 0.894) | 1.000 (0.990 - 1.000) | 567.87 (35.470 -9091.7) | 0.209 (0.125 - 0.350) | 2712.1 (157.06 - 46833.5) |
| Ciotti et al.(6) | 0.308 (0.170- 0.476) | 1.000 (0.715 - 1.000) | 7.500 (0.478 -117.57) | 0.717 (0.564 - 0.912) | 10.455 (0.570 - 191.78) |
| Kohmer et al.(7) | 0.290 (0.204 - 0.389) | 0.250 (0.169 - 0.347) | 0.387 (0.279 -0.536) | 2.840 (1.978 - 4.078) | 0.136 (0.073 - 0.255) |
| Kohmer et al.(7) | 0.320 (0.230 - 0.421) | 0.260 (0.177 - 0.357) | 0.432 (0.318 -0.589) | 2.615 (1.830 - 3.737) | 0.165 (0.090 - 0.305) |
| Kohmer et al.(7) | 0.180 (0.110 - 0.269) | 0.260 (0.177 - 0.357) | 0.243 (0.158 -0.375) | 3.154 (2.238 - 4.445) | 0.077 (0.039 - 0.152) |
| Kohmer et al.(7) | 0.370 (0.276 - 0.472) | 0.260 (0.177 - 0.357) | 0.500 (0.378 -0.662) | 2.423 (1.685 - 3.484) | 0.206 (0.113 - 0.377) |
| Linares et al. (22) | 0.157 (0.114 - 0.207) | 0.922 (0.881 - 0.951) | 2.000 (1.203 -3.324) | 0.915 (0.858 - 0.975) | 2.186 (1.239- 3.857) |
| Nalumansia et al.(23) | 0.700 (0.594 - 0.792) | 0.924 (0.874 - 0.959) | 9.262 (5.398 -15.891) | 0.325 (0.236 - 0.446) | 28.538 (13.848 - 58.814) |
| Pilarowski et al.(34) | 0.023 (0.008 - 0.053) | 0.960 (0.925 - 0.982) | 0.576 (0.196 -1.691) | 1.018 (0.984 - 1.052) | 0.566 (0.187 - 1.717) |
| Pilarowski et al.(34) | 0.556 (0.212- 0.863) | 0.503 (0.462 - 0.545) | 1.119 (0.620 -2.018) | 0.883 (0.423 - 1.841) | 1.267 (0.337 - 4.767) |
| Salvagno et al. (31) | 0.340 (0.288-0.394) | 0.994 (0.978 - 0.999) | 54.500 (13.575 -218.81) | 0.665 (0.614 - 0.719) | 82.007(20.035- 335.66) |
| Scohy et al. (24) | 0.378 (0.291-0.472) | 1.000 (0.916 - 1.000) | 32.608 (2.053 -517.87) | 0.628 (0.544 - 0.725) | 51.913 (3.118- 864.30) |
| Toptan et al.(25) | 0.500 (0.319 -0.681) | 1.000 (0.907 - 1.000) | 39.000 (2.432 -625.53) | 0.506 (0.359 - 0.714) | 77.000 (4.357 - 1360.8) |
| Torres et al.(26) | 0.060 (0.043-0.081) | 1.000 (0.994 - 1.000) | 77.000 (4.741 -1250.6) | 0.940 (0.922 - 0.959) | 81.905 (5.021- 1336.2) |
| Prince-Guerra et al.(35) | 0.525 (0.467-0.583) | 0.999 (0.997 - 1.000) | 409.57 (152.91 -1097.0) | 0.476 (0.422 - 0.536) | 861.29 (314.78 - 2356.6) |
| Courtellemont et al. (9) | 0.967 (0.918-0.991) | 1.000 (0.971 - 1.000) | 246.56 (15.502 -3921.4) | 0.037 (0.015 - 0.092) | 6658.3 (354.65 - 125004.7) |
| Courtellemont et al. (9) | 0.706 (0.525-0.849) | 1.000 (0.897 - 1.000) | 49.000 (3.100 -774.56) | 0.304 (0.183 - 0.506) | 161.00 (9.002 - 2879.3) |
| Cerutti et al. (36) | 0.706 (0.612- 0.790) | 1.000 (0.983 - 1.000) | 312.82(19.576 - 4998.8) | 0.296 (0.222 - 0.395) | 1056.4 (63.918 - 17459.1) |

**Table 2.** Sub analysis of sensitivity and specificity for nasopharyngeal swab with 95% confidence interval.

**Figure 3.** A forest plot showing the estimates for sensitivity (A) and specificity (B) for nasopharyngeal swab.

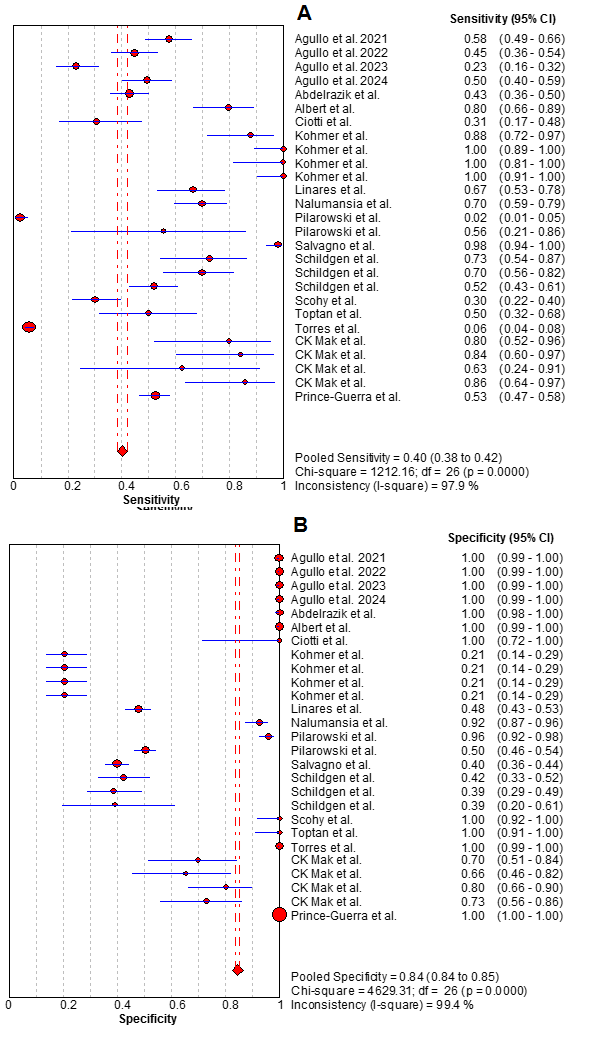

**Table 3.** Sub analysis of sensitivity and specificity for throat washing and broncho-alveolar fluids with 95% confidence interval.

| Study | Sensitivity | Specificity | Positive LR | Negative LR | DOR |
| --- | --- | --- | --- | --- | --- |
| Schildgen et al. (32) | 0.329 (0.223- 0.449) | 0.877 (0.779- 0.942) | 2.667 (1.332 - 5.338) | 0.766 (0.638 - 0.919) | 3.483(1.486- 8.162) |
| Schildgen et al. (32) | 0.500 (0.381- 0.619) | 0.781 (0.669- 0.869) | 2.281 (1.399 - 3.721) | 0.640 (0.495- 0.829) | 3.563 (1.738 - 7.302) |
| Schildgen et al. (32) | 0.877 (0.779- 0.942) | 0.795 (0.684- 0.880) | 4.267 (2.696- 6.753) | 0.155(0.083 - 0.289) | 27.496(11.184- 67.599) |
| CK Mak et al. (18) | 0.400 (0.257- 0.557) | 1.000 (0.921- 1.000) | 37.000 (2.297 - 595.89) | 0.604 (0.476- 0.768) | 61.218 (3.546- 1056.9) |

A

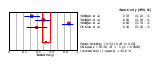

B

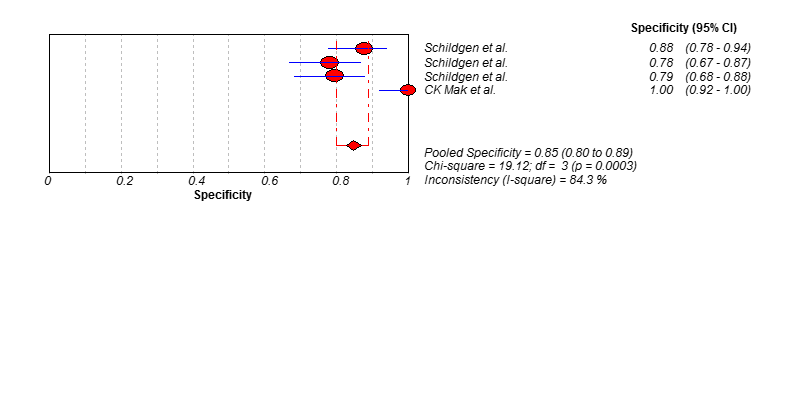

**Figure 4.** A forest plot showing the estimates for sensitivity (A) and specificity (B) for Throat washing and Bronchoalveolar fluids.

**Table 4.** Sub analysis of sensitivity and specificity for finger-stick whole-blood with 95% confidence interval.

| Study | Sensitivity | Specificity | Positive LR | Negative LR | DOR |
| --- | --- | --- | --- | --- | --- |
| Pere et al. (10) | 0.958 (0.857- 0.995) | 0.981 (0.897 - 1.000) | 49.833 (7.147- 347.45) | 0.042 (0.011- 0.165) | 1173.0 (102.92- 13368.3) |
| Pere et al. (10) | 0.917 (0.800- 0.977) | 0.865 (0.742 - 0.944) | 6.810 (3.401 - 13.636) | 0.096 (0.037- 0.248) | 70.714 (19.333- 258.66) |
| Pere et al. (10) | 0.923 (0.749- 0.991) | 1.000 (0.858 - 1.000) | 45.370 (2.910- 707.30) | 0.094 (0.029- 0.308) | 480.20 (21.903- 10527.9) |
| Pere et al. (10) | 0.979 (0.889- 0.999) | 0.981 (0.897- 1.000) | 50.917 (7.306 - 354.84) | 0.021 (0.003- 0.148) | 2397.0 (145.76- 39418.0) |
| Pere et al. (10) | 0.915 (0.796- 0.976) | 0.846 (0.719 - 0.931) | 5.947 (3.125 - 11.316) | 0.101 (0.039- 0.259) | 59.125 (16.576- 210.89) |

A	B

a
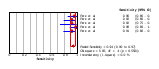
b
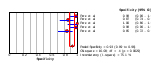

**Figure 5.** A forest plot showing the estimates for sensitivity (A) and specificity (B) for Finger-stick whole-blood.

**Table 5.** Sub analysis of sensitivity and specificity for Symptomatic patients with 95% confidence interval.

| Study | Sensitivity | Specificity | Positive LR | Negative LR | DOR |
| --- | --- | --- | --- | --- | --- |
| Agullo et al. (27) | 0.147 (0.111- 0.188) | 1.000 (0.923- 1.000) | 13.871 (0.870 - 221.04) | 0.862 (0.817- 0.908) | 16.099 (0.977- 265.36) |
| Agullo et al. (27) | 0.083 (0.054- 0.120) | 1.000 (0.944- 1.000) | 10.941 (0.675- 177.41) | 0.923 (0.886- 0.961) | 11.854 (0.712- 197.28) |
| Agullo et al. (27) | 0.166 (0.127- 0.210) | 1.000 (0.905- 1.000) | 12.667 (0.799 - 200.93) | 0.844 (0.795- 0.897) | 15.000 (0.908- 247.93) |
| Prince-Guerra et al. (35) | 0.642 (0.566- 0.713) | 1.000 (0.994- 1.000) | 836.18 (52.243- 13383.6) | 0.359 (0.295 - 0.437) | 2329.0 (143.07- 37912.1) |
| Scohy et al. (24) | 0.291 (0.198 - 0.399) | 0.395 (0.292- 0.507) | 0.481 (0.332- 0.697) | 1.794 (1.337- 2.408) | 0.268 (0.142- 0.506) |
| Schildgen et al. (32) | 0.304 (0.132- 0.529) | 0.783 (0.563- 0.925) | 1.400 (0.519- 3.773) | 0.889 (0.629- 1.256) | 1.575 (0.416 - 5.959) |
| Schildgen et al. (32) | 0.391 (0.197- 0.615) | 0.826 (0.612- 0.950) | 2.250 (0.806- 6.279) | 0.737 (0.505- 1.075) | 3.054 (0.780 - 11.959) |
| Schildgen et al. (32) | 1.000 (0.852- 1.000) | 0.087 (0.011- 0.280) | 1.093 (0.942 - 1.268) | 0.200 (0.010- 3.950) | 5.465 (0.248 - 120.37) |

**Figure 6.** A forest plot showing the estimates for sensitivity (A) and specificity (B) for Symptomatic patients.

A

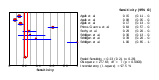

B

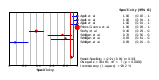

**Table 6.** Sub analysis of sensitivity and specificity for asymptomatic patients with 95% confidence interval.

| Study | Sensitivity | Specificity | Positive LR | Negative LR | DOR |
| --- | --- | --- | --- | --- | --- |
| Agullo et al. (27) | 0.034 (0.015- 0.065) | 1.000 (0.872 - 1.000) | 1.992 (0.118-33.587) | 0.982 (0.929- 1.038) | 2.028 (0.114 -36.111) |
| Agullo et al. (27) | 0.014 (0.003- 0.040) | 1.000 (0.881 - 1.000) | 0.972 (0.051-18.363) | 1.000 (0.952- 1.051) | 0.972 (0.049-19.288) |
| Agullo et al. (27) | 0.041 (0.019- 0.076) | 1.000 (0.858 - 1.000) | 2.140 (0.128-35.668) | 0.977 (0.918- 1.040) | 2.191 (0.124-38.806) |
| Prince-Guerra et al. (35) | 0.358 (0.273- 0.449) | 0.998 (0.996 - 1.000) | 220.80 (80.629-604.68) | 0.643 (0.564- 0.734) | 343.23 (120.37-978.66) |
| Scohy et al. (24) | 0.089 (0.025- 0.212) | 0.689 (0.534 - 0.818) | 0.286 (0.102-0.802) | 1.323 (1.065- 1.642) | 0.216 (0.065-0.721) |
| Courtellemont et al. (9) | 1.000 (0.858- 1.000) | 0.886 (0.733 - 0.968) | 7.840 (3.297-18.640) | 0.023 (0.001- 0.356) | 343.00 (17.613-6679.5) |
| Schildgen et al. (32) | 0.296 (0.138- 0.502) | 0.926 (0.757- 0.991) | 4.000 (0.934-17.134) | 0.760 (0.582- 0.993) | 5.263 (1.000-27.690) |
| Schildgen et al. (32) | 0.370 (0.194- 0.576) | 0.704 (0.498- 0.862) | 1.250 (0.584-2.677) | 0.895 (0.613 - 1.307) | 1.397 (0.448- 4.355) |
| Schildgen et al. (32) | 0.852 (0.663 - 0.958) | 0.148 (0.042- 0.337) | 1.000 (0.801-1.249) | 1.000 (0.278 - 3.594) | 1.000 (0.223- 4.489) |

A

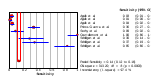

B

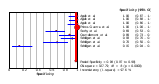

**Figure 7.** A forest plot showing the estimates for sensitivity (A) and specificity (B) for asymptomatic patients.
